# LLM-Based Annotation and Token-Augmented Modeling for Emotional Tone Classification in Online Cancer Peer-Support Posts

**DOI:** 10.64898/2026.01.27.26344999

**Authors:** Shuo Xu, Zhongyan Wang, Hailiang Wang, Zhanyi Ding, Yue Zou, Yuchen Cao

**Affiliations:** Computer Science and Engineering Department, University of California San Diego, La Jolla, CA, USA; Center for Data Science, New York University, New York, NY, USA; College of Computing, Georgia Institute of Technology, Atlanta, GA, USA; Fu Foundation School of Engineering and Applied Science, Columbia University, New York, NY, USA; Khoury college of computer science, Northeastern University, Seattle, WA, USA

## Abstract

Online cancer peer-support communities generate large volumes of patient-authored and caregiver-authored text that may reflect distress, coping, and informational needs. Automated emotional tone classification could support scalable monitoring, but supervised modeling depends on label quality and may benefit from explicit context features. Using the *Mental Health Insights: Vulnerable Cancer Survivors & Caregivers* dataset, we compared five model families (TF–IDF Logistic Regression, Random Forest, LightGBM, GRU, and fine-tuned ALBERT) on a three-class target (Negative/Neutral/Positive) derived from four original categories. We introduced two extensions: (i) LLM-based annotation to generate parallel “AI labels” and (ii) token-based augmentation that prepends LLM-extracted structured variables (reporter role and cancer type) to the post text. Models were trained with a 60/20/20 stratified train/validation/test split, with hyperparameters selected on validation data only. Test performance was summarized using weighted F1 and macro one-vs-rest AUC with bootstrap confidence intervals, with paired comparisons based on McNemar tests and false discovery rate adjustment. The LLM annotator produced substantial redistribution in the four-class label space, shifting prevalence toward *very negative* relative to the original labels; the shift persisted but attenuated after collapsing to three classes. Across all model families, token augmentation improved held-out performance, with the largest gains for GRU and consistent improvements for ALBERT. Augmentation also reduced polarity-reversing errors (Nega-↔ tive Positive) for ALBERT, while adjacent errors (Negative ↔ Neutral) remained the dominant residual failure mode. These results indicate that LLM-based supervision can introduce systematic measurement shifts that require auditing, yet LLM-extracted context incorporated via simple token augmentation provides a pragmatic, model-agnostic mechanism to improve downstream emotional tone classification for supportive oncology decision support.

**Author summary:** We studied how to better monitor emotional tone in posts from online cancer peer-support communities, where patients and caregivers share experiences that may signal distress, coping, or unmet needs. Automated classification could help organizations and moderators identify when additional support may be needed, but these systems depend on the quality of the labels used for training and may miss clinical context. Using a public dataset of cancer survivor and caregiver posts, we trained and compared several machine-learning and deep-learning models to classify each post as negative, neutral, or positive. We tested two practical improvements. First, we used a large language model to generate an additional set of “AI labels” and examined how these differed from the original categories. Second, we extracted simple context information—whether the writer was a patient or caregiver and what cancer type was mentioned—and added this context to the text before model training. We found that adding context consistently improved performance across model types. However, the AI-generated labels shifted class distributions, indicating that automated labeling can introduce systematic changes that should be audited. Overall, simple context extraction can make emotional tone monitoring more accurate and useful for supportive oncology decision support.

## 1 Introduction

Cancer survivorship and treatment are frequently accompanied by substantial psychosocial burden, affecting both patients and informal caregivers. As survivorship populations grow, supportive care and distress monitoring have become central to quality oncology care, yet practical screening and follow-up remain resource constrained [1, 2]. In parallel, many individuals turn to online peer-support communities to seek information, disclose symptoms, and process uncertainty. These posts constitute a form of patient-generated health data (PGHD) that can complement traditional clinical touchpoints, but their volume and heterogeneity make manual review difficult to sustain at scale [3, 4].

Natural language processing (NLP) offers a pathway to summarize and triage supportive-care needs from non-clinical text, and sentiment/emotion modeling has become a common first step for identifying distress-related language in social media and other user-generated content [5–7]. However, the operational use of sentiment classifiers in health contexts faces two persistent challenges. First, emotional expression in cancer discourse is nuanced and often mixed, with posts blending informational content, coping narratives, and acute distress. This can lead to ambiguous boundaries between adjacent affective categories (e.g., negative vs. neutral) and to class imbalance where positive coping language is relatively rare. Second, developing supervised models depends critically on labels that may be noisy, inconsistent across raters, and sensitive to the annotation rubric—issues that can materially affect downstream learnability and model evaluation [8, 9].

Recent work has proposed using large language models (LLMs) as scalable annotators for text classification tasks, sometimes achieving strong agreement with human judgments while reducing labeling time and cost [10]. At the same time, LLM-based labeling can introduce systematic shifts in decision boundaries, particularly for subjective constructs such as emotional intensity, and may exhibit overconfidence or error modes that are not immediately evident without auditing [11, 12]. For mental-health-adjacent content, such shifts are especially consequential: a more conservative rater may inflate negative prevalence, alter apparent community needs, and bias models trained or evaluated on those labels. Thus, when LLMs are used to generate labels, it is important not only to report predictive performance but also to characterize label distribution changes and to study how downstream models respond under controlled evaluation.

In this work, we investigate emotional tone classification in online cancer peer-support posts using the *Mental Health Insights: Vulnerable Cancer Survivors & Caregivers* dataset [13]. The dataset provides author-reported emotional tone labels across four categories (very negative, negative, neutral, positive). We extend this setting by applying an LLM annotator to produce (i) a parallel set of emotional tone labels (AI labels) and (ii) lightweight structured variables extracted from the same text (reporter role and cancer type). This design enables a focused methodological study of two questions: how LLM labeling reallocates emotional tone relative to existing labels, and whether LLM-derived structured context can be incorporated to improve downstream classification.

Methodologically, we compare a range of commonly used model families for text classification, spanning TF–IDF-based classical baselines and neural encoders, including a fine-tuned transformer (ALBERT) [14, 15]. We further evaluate a simple, model-agnostic mechanism for leveraging LLM-extracted variables: token-based augmentation, in which deterministic prefix tokens (e.g., `ROLE PATIENT, CANCER LUNG`)are prepended to the post text. This approach allows structured context to be consumed by both sparse vectorizers and neural tokenizers without introducing multi-branch architectures, providing a pragmatic template for integrating upstream information extraction with downstream screening.

Our contributions are threefold. First, we provide an empirical characterization of human versus LLM label distributions under both the original four-class scheme and a three-class collapse used for modeling, highlighting the extent and nature of labeling shift. Second, we quantify the effect of token-based augmentation across multiple model families under a controlled train/validation/test design, including uncertainty estimates and paired statistical comparisons. Third, we analyze error structure with attention to ordinal distance (adjacent vs. polarity-reversing errors), emphasizing practical implications for decision-support deployment in supportive oncology settings. Collectively, these results aim to clarify when LLMs function as helpful annotators and context extractors, when they induce systematic artifacts, and how simple augmentation strategies can improve robustness and minority-class behavior in emotionally salient health-related text.

## 2 Methods

We conducted a comparative modeling study to classify the emotional tone of posts from online cancer peer-support communities. The overall experimental framework (data source, train/validation/test splitting, preprocessing decisions by model family, model families, and evaluation procedures) followed the same pipeline as our related work. The substantive extensions in this study are that (i) prediction targets were derived from an LLM-based annotation procedure (AI labels) and (ii) selected LLM-derived structured variables were incorporated as additional model inputs via token-based augmentation. Hyperparameter selection was performed using a held-out validation set only, and final performance was reported on an untouched test set.

### Computational environment

All experiments were implemented in Python. Data manipulation used NumPy and Pandas. Classical machine learning components (data partitioning, TF–IDF construction, model fitting, and hyperparameter search) were implemented with `scikit-learn`[16]. Gradient boosting was implemented using `LightGBM`[17]. Neural network models were developed in `PyTorch`, and transformer fine-tuning used the Hugging Face `transformers`library with the pretrained `albert-base-v2`checkpoint [15]. Text standardization utilities used Python regular expressions and NLTK resources [18]. Training progress was tracked with `tqdm`. When available, CUDA acceleration was used for neural model training. The GRU architecture followed the standard gated recurrent unit formulation [19].

### Dataset

We used the *Mental Health Insights: Vulnerable Cancer Survivors & Caregivers* dataset [13], a de-identified corpus of user-authored posts from online cancer support communities. Each instance consists of a free-text post and an author-provided emotional tone label spanning four categories: `very negative, negative, neutral`, and `positive`. In this study, we additionally applied an LLM annotator to the raw post text to generate (i) a parallel set of emotional tone labels (AI labels) and (ii) lightweight structured variables used for text augmentation (reporter role and cancer type).

#### Four-class annotation vs. three-class modeling target

Although the source dataset is four-class, our downstream predictive experiments use a three-class target obtained by collapsing VERY NEGATIVEand `NEGATIVE`into a single *Negative* class, while retaining `NEUTRAL`and `POSITIVE`as distinct classes. This reduction follows common practice in affect modeling for health-related social media where the boundary among negative subclasses is often less stable and less actionable for downstream use. We derived the three-class target consistently for both the original labels and the AI labels using the mapping

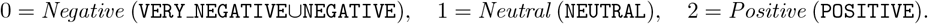

Unless otherwise stated, all reported predictive results use this three-class target. The original four-class labels and the LLM-produced four-class probabilities were retained for descriptive analyses of labeling behavior (Section 3).

### Train/validation/test splitting

We used a two-stage holdout design. First, 20% of the dataset was reserved as the final test set. The remaining 80% was split into training and validation subsets, producing an overall 60/20/20 train/validation/test split. All splits were generated with a fixed seed (`random state=42`). To preserve class proportions under imbalance, splits were stratified by the three-class target. The validation set was used for hyperparameter selection; the test set was used only for final reporting.

### Text processing and representations

#### Model-family preprocessing

Preprocessing was aligned to common practice for different model families. For sparse lexical baselines (Logistic Regression, Random Forest, LightGBM), models were trained on TF–IDF representations derived from the input text with built-in English stopword removal and without additional lemmatization. For neural models (GRU and ALBERT), we applied a lightweight standardization pipeline: text was lowercased; URLs, user mentions, and hashtags were replaced with generic placeholders via regular expressions; non-alphanumeric characters were removed; and whitespace was normalized. This approach reduces noisy artifacts while preserving contextual cues for neural tokenization.

#### Feature construction

For classical models, TF–IDF vectors were built using up to 10,000 features, including unigrams and bigrams (`ngram range=(1,2)`). For neural models, ALBERT used `AlbertTokenizer`with a maximum sequence length of 200 tokens, and the GRU model used a vocabulary of the top 10,000 tokens with sequences padded/truncated to length 200.

### LLM-based labeling and structured variable extraction

#### Annotator and protocol

We used an LLM (`gpt-4o-mini`) as an automated annotator to extract emotional tone labels and a small set of auxiliary variables from each post. The annotator was instructed to avoid clinical diagnosis or medical advice and to base outputs strictly on the provided text. Posts were provided in batched JSON format (20 posts per batch), and the model returned a JSON array of structured outputs in the same order. To maximize completeness under occasional formatting failures, we implemented a recursive split-and-retry strategy in which a failed batch was subdivided until valid JSON was recovered or a single-post failure remained. Outputs were written with checkpointing to enable safe resumption.

#### AI labels (four-class probabilities and discrete label)

For each post, the annotator produced emo probs, a probability distribution over four emotional tone categories: `VERY NEGATIVE, NEGATIVE, NEUTRAL,` and `POSITIVE`, with probabilities constrained to be non-negative and to sum to 1 (within rounding tolerance). A discrete four-class AI label was defined as the argmax category, using the mapping 0=VERY NEGATIVE, 1=`NEGATIVE`, 2=`NEUTRAL`, 3=`POSITIVE`. For the three-class predictive task, the AI label was then collapsed deterministically to the three-class mapping in Section 2.

#### Auxiliary variables for augmentation

In addition to emotional tone, the annotator extracted two auxiliary variables used for augmentation: (i) reporter role (`PATIENT, CAREGIVER, UNCLEAR`) and (ii) cancer type (`BRAIN, COLON, LIVER, LEUKEMIA, LUNG, OTHER,UNKNOWN`), along with an associated confidence score (0–1). Evidence strings returned by the annotator were retained for qualitative auditing but were not used as model inputs. Importantly, these structured variables are extracted from the same post text and therefore do not introduce external information; rather, they emulate an upstream information-extraction step that produces coarse context features to support downstream screening.

### Modeling conditions and token-based augmentation

We fit two modeling conditions that differed only in the input text field consumed by the learning algorithms.

#### Regular (text-only) models

In the *regular* condition, each model consumed the original post text (posts) without additional structured variables.

#### Augmented models (reporter role and cancer type tokens)

In the *augmented* condition, we prepended deterministic tokens encoding the LLM-derived reporter role and cancer type to the post text to form an augmented input string (`posts aug`). This design allows structured variables to be consumed by both TF–IDF vectorizers and neural tokenizers without requiring multi-branch architectures.

#### Token definitions and construction

Reporter role was encoded as `ROLE <ROLE>`(e.g., `ROLE PATIENT`). Values outside `{PATIENT, CAREGIVER, UNCLEAR}`were mapped to `ROLE UNCLEAR`. Cancer type was encoded as `CANCER <TYPE>`(e.g., `CANCER LUNG`). Values outside the allowed set were mapped to `CANCER UNKNOWN`. The final augmented input was constructed as

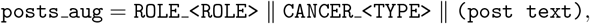

where ∥ denotes string concatenation with a single separating space. All models in the augmented condition consumed `posts aug`. For classical baselines, TF–IDF vectorization was applied directly to `posts aug`, so covariate tokens entered the sparse vocabulary as additional lexical features. For neural models, `posts aug`was passed through the same tokenization pipeline; covariate tokens therefore contributed as prefix tokens within the encoded sequence.

### Models and hyperparameter optimization

We evaluated five model families: multinomial Logistic Regression, Random Forest, LightGBM, a GRU sequence model, and a fine-tuned ALBERT transformer. To mitigate class imbalance, we used class-weighted objectives or balanced weighting options where supported. Classical models were tuned using grid search over standard regularization and tree/boosting parameters. Neural models (GRU and ALBERT) were tuned using random search over learning rate, dropout, and training epochs. For all model families, the primary selection criterion was validation weighted F1-score computed on the three-class target. The full search spaces and selected hyperparameters are summarized in Table 1.

**Table 1.**
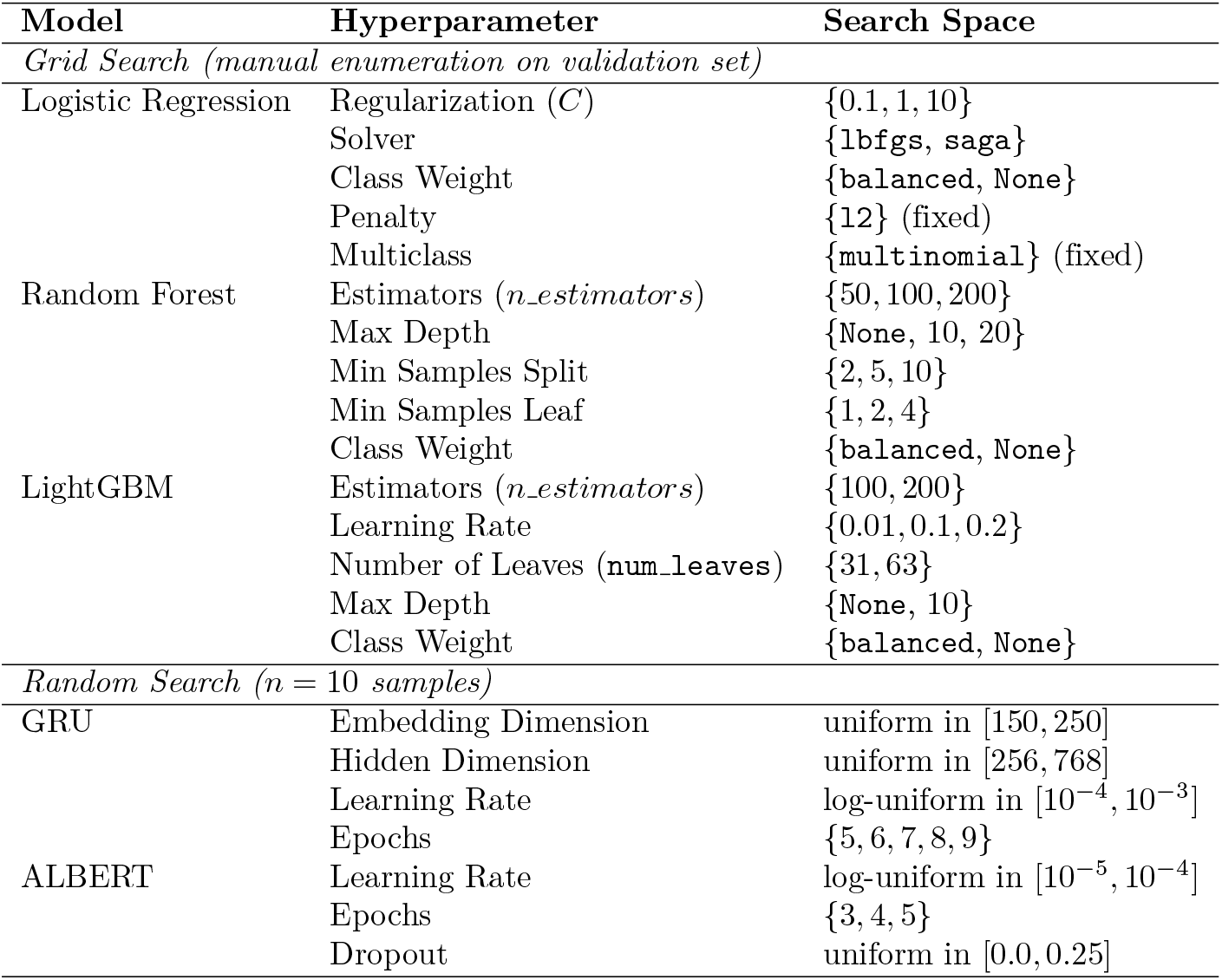
Hyperparameter Search Spaces for ML and DL Models.

### Evaluation and statistical comparison

We reported accuracy and class-wise precision, recall, and F1-score, using weighted F1 as the primary aggregate metric. Discriminative ability was additionally summarized using macro-averaged one-vs-rest (OvR) ROC AUC.

To quantify uncertainty in aggregate performance, 95% confidence intervals for weighted F1 and macro OvR AUC were computed using nonparametric bootstrapping with 2,000 resamples of the held-out test set. For each resample, metrics were recomputed and percentile-based intervals were formed from the empirical bootstrap distribution.

For paired comparisons between models evaluated on the same test instances, we applied McNemar’s test to per-instance correctness using the continuity-corrected asymptotic formulation. To control multiplicity across pairwise tests within each modeling condition, p-values were adjusted using the Benjamini–Hochberg false discovery rate procedure, with significance assessed at *α* = 0.05.

### Ordinal-aware error analysis

Although the downstream task uses a three-class target, emotional tone remains conceptually ordered from negative to positive. We therefore examined confusion matrices with attention to the distance of misclassification, distinguishing adjacent-category confusions from distant-category confusions to identify error modes with greater potential operational consequences, such as under-identification of positive coping language or misclassification of negative narratives as neutral.

## 3 Results

We report (i) overall held-out test performance under *regular* (text-only) versus *augmented* (role+cancer-type tokens) inputs, (ii) class-specific effects of augmentation for the best-performing model family, and (iii) paired statistical comparisons based on per-instance correctness. Unless otherwise noted, results correspond to the three-class modeling target (Negative/Neutral/Positive) derived from the four-class label space as described in Section 2.

### Human vs. LLM Label Distributions (4-class and Collapsed 3-class)

To characterize how the LLM annotator reallocated emotional tone relative to the dataset’s original labels, we compared class prevalence under the four-class scheme and under the downstream three-class collapse (Negative vs. Neutral vs. Positive). Substantial distributional shift was observed in the four-class comparison. Under human labels, the corpus was dominated by *neutral* (42.1%) and *negative* (39.6%), with *very negative* (11.1%) and *positive* (7.2%) as minority categories. In contrast, the LLM labeling reassigned nearly half of all posts to *very negative* (46.3%), while reducing the prevalence of *negative* (24.9%), *neutral* (24.0%), and *positive* (4.7%). This divergence was statistically pronounced (chi-square = 13022.21, *p <* 0.001) and reflected a moderate distributional distance (Jensen–Shannon divergence = 0.1155), indicating that the LLM applied a substantially different decision boundary for intensity between *very negative* and the mid-spectrum classes.

When collapsing *very negative* and *negative* into a single *Negative* class (the target used for three-class modeling), the shift remained but was attenuated. The human-label distribution was 50.7% Negative, 42.1% Neutral, and 7.2% Positive, whereas the LLM distribution became 71.2% Negative, 24.0% Neutral, and 4.7% Positive. The collapsed comparison was still highly significant (chi-square = 1757.97, *p <* 0.001) but exhibited a smaller Jensen–Shannon divergence (= 0.0324), suggesting that most disagreement arises from how the LLM partitions *very negative* versus *negative* and from a systematic tendency to label informational or mixed-tone posts as negative rather than neutral. Practically, this implies that analyses relying on the LLM labels will be more conservative with respect to distress detection (higher negative prevalence), while potentially under-representing neutral and positive content; downstream modeling and interpretation should therefore treat the AI labels as a methodological artifact and evaluate robustness to this labeling shift.

### Overall test performance under regular vs. augmented inputs

Table 2 summarizes test-set performance for each model family under two input conditions: (Panel A) *regular* text-only inputs and (Panel B) *augmented* inputs that prepend LLM-derived `ROLE_<ROLE>`and `CANCER_<TYPE>`tokens to each post. Across both panels, we report weighted-average precision and recall, along with weighted F1 and macro one-vs-rest (OvR) AUC with 95% bootstrap confidence intervals.

**Table 2.** Overall Model Performance Summary on Test Set.

| Model | Precision | Recall | Weighted F1 (95% CI) | Macro AUC (95% CI) |
| --- | --- | --- | --- | --- |
| *Panel A. Regular version* |  |  |  |  |
| Logistic Regression | 0.8588 | 0.8615 | 0.8457 [0.8283, 0.8624] | 0.8992 [0.8823, 0.9146] |
| Random Forest | 0.8495 | 0.8466 | 0.8406 [0.8237, 0.8561] | 0.8684 [0.8476, 0.8885] |
| LightGBM | 0.8480 | 0.8567 | 0.8457 [0.8286, 0.8624] | 0.8761 [0.8578, 0.8945] |
| GRU | 0.8072 | 0.7975 | 0.8013 [0.7835, 0.8181] | 0.8051 [0.7800, 0.8295] |
| ALBERT | 0.8610 | 0.8393 | 0.8476 [0.8328, 0.8623] | 0.9177 [0.9029, 0.9306] |
| *Panel B. Augmented version* |  |  |  |  |
| Logistic Regression | 0.8629 | 0.8663 | 0.8513 [0.8339, 0.8682] | 0.9059 [0.8895, 0.9215] |
| Random Forest | 0.8518 | 0.8581 | 0.8477 [0.8310, 0.8644] | 0.8807 [0.8604, 0.8997] |
| LightGBM | 0.8485 | 0.8499 | 0.8472 [0.8317, 0.8635] | 0.8870 [0.8678, 0.9046] |
| GRU | 0.8290 | 0.8201 | 0.8231 [0.8068, 0.8386] | 0.8407 [0.8183, 0.8620] |
| ALBERT | 0.8654 | 0.8562 | 0.8598 [0.8454, 0.8748] | 0.9284 [0.9164, 0.9399] |

Precision and Recall correspond to weighted-average values on the test set. Weighted F1 and Macro AUC are reported with 95% confidence intervals.

Across all five model families, augmentation improved aggregate performance on the held-out test set. Weighted F1 increased from Panel A to Panel B for Logistic Regression (0.8457 →0.8513; Δ = +0.0056), Random Forest (0.8406 →0.8477; Δ = +0.0071), LightGBM (0.8457 →0.8472; Δ = +0.0015), GRU (0.8013 →0.8231; Δ = +0.0218), and ALBERT (0.8476 →0.8598; Δ = +0.0122). Macro AUC increased in parallel: Logistic Regression (0.8992 →0.9059; Δ = +0.0067), Random Forest (0.8684 →0.8807; Δ = +0.0123),LightGBM (0.8761 →0.8870; Δ = +0.0109), GRU (0.8051 →0.8407; Δ = +0.0356), and ALBERT (0.9177 →0.9284; Δ = +0.0107). The uniform direction of these gains supports the central premise that lightweight structured context derived from the same post text (who is speaking and which cancer type is referenced) provides complementary signal that can be exploited by both sparse lexical models and neural encoders. Notably, GRU exhibited the largest absolute improvements on both weighted F1 and macro AUC, consistent with high-salience prefix tokens providing additional guidance to a lower-capacity sequential model.

Model ordering remained broadly consistent across conditions. ALBERT achieved the highest point estimates of weighted F1 and macro AUC in both panels (Panel A: weighted F1=0.8476; macro AUC=0.9177; Panel B: weighted F1=0.8598; macro AUC=0.9284), indicating strong discriminative performance even prior to augmentation and additional gains after introducing context tokens. Differences among the top-performing approaches were modest in weighted F1, with overlapping confidence intervals among Logistic Regression, LightGBM, and ALBERT in both panels, suggesting that multiple architectures achieved comparable aggregate classification quality under this three-class target. Macro AUC more clearly separated ALBERT from classical baselines, indicating stronger global discrimination across classes.

### Class-specific effects of augmentation in the best-performing model (ALBERT)

To localize the source of aggregate gains, we examined class-specific precision, recall, and F1 for ALBERT under the regular and augmented conditions (Table 3). Class-wise results indicate that augmentation reshaped error patterns rather than uniformly inflating performance.

**Table 3.**
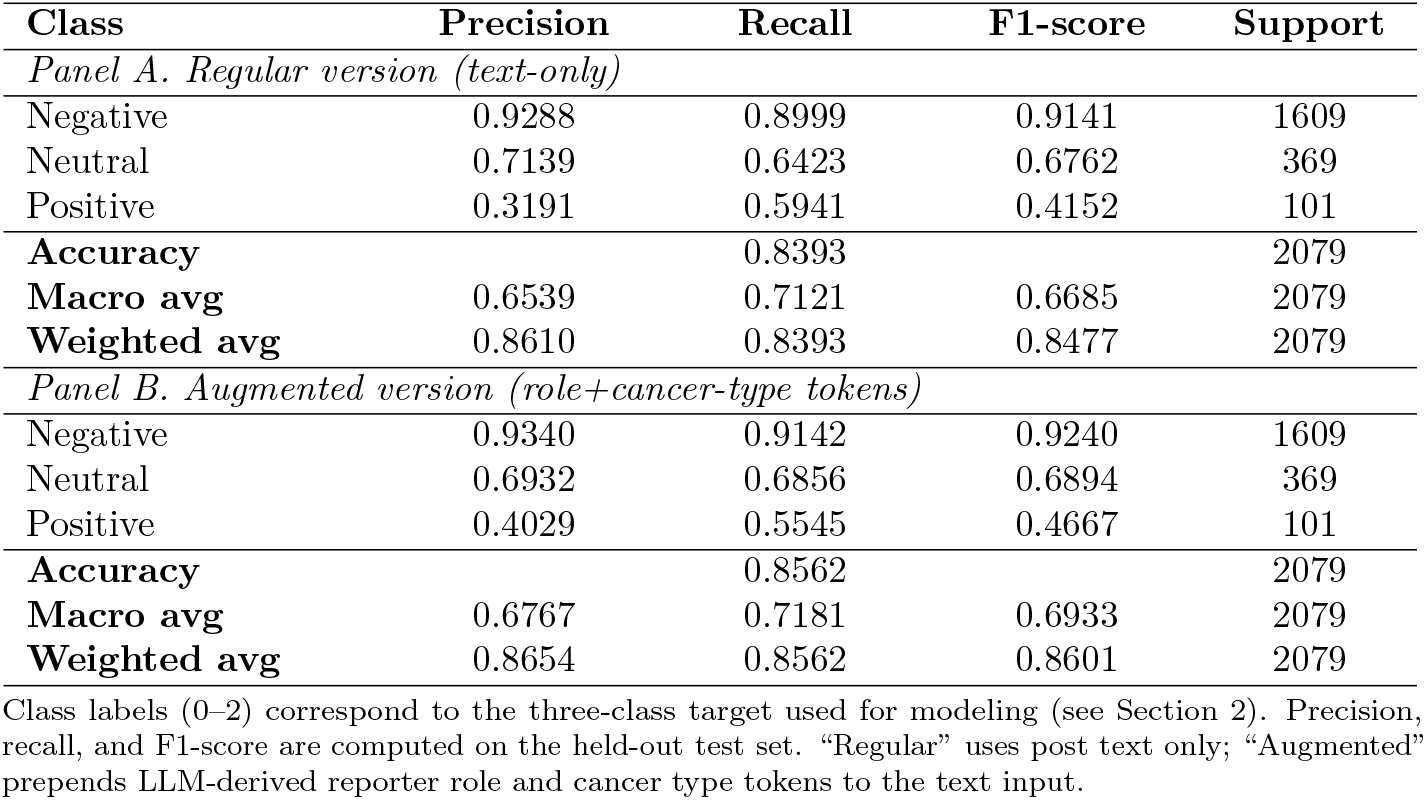
ALBERT Class-Specific Performance on the Test Set (Regular vs. Augmented Inputs)

| Class | Precision | Recall | F1-score | Support |
| --- | --- | --- | --- | --- |
| *Panel A. Regular version (text-only)* |  |  |  |  |
| Negative | 0.9288 | 0.8999 | 0.9141 | 1609 |
| Neutral | 0.7139 | 0.6423 | 0.6762 | 369 |
| Positive | 0.3191 | 0.5941 | 0.4152 | 101 |
| **Accuracy** |  | 0.8393 |  | 2079 |
| **Macro avg** | 0.6539 | 0.7121 | 0.6685 | 2079 |
| **Weighted avg** | 0.8610 | 0.8393 | 0.8477 | 2079 |
| *Panel B. Augmented version (role+cancer-type tokens)* |  |  |  |  |
| Negative | 0.9340 | 0.9142 | 0.9240 | 1609 |
| Neutral | 0.6932 | 0.6856 | 0.6894 | 369 |
| Positive | 0.4029 | 0.5545 | 0.4667 | 101 |
| **Accuracy** |  | 0.8562 |  | 2079 |
| **Macro avg** | 0.6767 | 0.7181 | 0.6933 | 2079 |
| **Weighted avg** | 0.8654 | 0.8562 | 0.8601 | 2079 |

Class labels (0–2) correspond to the three-class target used for modeling (see Section 2). Precision, recall, and F1-score are computed on the held-out test set. “Regular” uses post text only; “Augmented” prepends LLM-derived reporter role and cancer type tokens to the text input.

For the majority class (Negative; *n* = 1609), augmentation produced consistent improvement: precision increased from 0.9288 to 0.9340, recall from 0.8999 to 0.9142, and F1 from 0.9141 to 0.9240. This indicates reduced confusion of clearly negative posts with neutral/positive content after adding role and cancer-type context.

For the intermediate class (Neutral; *n* = 369), augmentation primarily improved sensitivity: recall increased from 0.6423 to 0.6856 and F1 increased from 0.6762 to 0.6894, while precision decreased modestly from 0.7139 to 0.6932. This pattern suggests that the added context enabled the model to recover additional neutral instances, at the cost of slightly more neutral assignments among non-neutral posts.

For the smallest class (Positive; *n* = 101), augmentation yielded the largest F1 gain (0.4152 →0.4667). This improvement was driven chiefly by a substantial precision increase (0.3191 →0.4029), while recall decreased slightly (0.5941 →0.5545), indicating improved specificity for positive coping language and fewer false-positive positive predictions. In aggregate, these shifts align with the observed increases in test accuracy (0.8393 →0.8562) and weighted F1 (0.8476 →0.8598), and suggest that deterministic context tokens can meaningfully alter decision boundaries for minority and boundary-sensitive classes rather than merely reinforcing already-strong majority-class performance.

### Paired statistical comparisons

To assess whether observed differences reflected systematic changes in per-instance correctness, we conducted continuity-corrected McNemar tests for all model pairs within each condition, with Benjamini–Hochberg adjustment to control the false discovery rate at *α* = 0.05 (Section 2). Consistent with the aggregate metrics, the most pronounced and statistically reliable differences involved the GRU baseline. In the regular condition, GRU was significantly outperformed by the classical TF–IDF baselines after adjustment, while differences among Logistic Regression, Random Forest, LightGBM, and ALBERT were generally not statistically distinguishable given their close aggregate performance. In the augmented condition, paired comparisons continued to favor non-GRU models, but the number of significant adjusted differences decreased, consistent with augmentation lifting performance across model families and narrowing gaps in per-instance correctness.

Taken together, the results indicate that (i) token-based incorporation of LLM-derived context improves performance consistently across model classes, and (ii) transformer fine-tuning (ALBERT) provides the strongest overall discrimination, particularly in macro AUC, while (iii) augmentation yields its most practically relevant gains by improving minority-class behavior and reducing specific confusions rather than producing uniform improvements across all classes.

### Error Analysis: Ordinal error structure and the impact of token augmentation

To complement aggregate metrics, we analyzed error structure on the held-out test set using the *regular* (text-only) and *augmented* (role+cancer-type tokens) prediction files. Because the three-class target is ordinal, we distinguished *adjacent* errors (distance = 1 on the label scale) from *distant* errors (distance = 2, i.e., Negative ↔ Positive), which are operationally more consequential.

Augmentation reduced the total number of incorrect predictions for most model families, with the largest absolute gains observed for the GRU and the best-performing transformer. Specifically, GRU errors decreased from 381 to 328 (net −53), reflecting 99 instances corrected by augmentation and 46 newly misclassified. ALBERT errors decreased from 334 to 299 (net −35), corresponding to 72 corrected instances and 37 newly incorrect. Classical models exhibited smaller shifts (Logistic Regression: net −16; Random Forest: net −24; LightGBM: net +3), consistent with augmentation acting as a modest but generally helpful signal that is most beneficial for lower-capacity sequential modeling and contextual encoders.

For ALBERT, augmentation yielded a meaningful reduction in the most severe ordinal errors. The number of distance-2 errors (Negative ↔ Positive) decreased from 107/334 (32.0%) in the regular setting to 71/299 (23.7%) in the augmented setting, indicating that the added context tokens helped the model avoid polarity-reversing mistakes. This improvement was driven primarily by fewer Negative →Positive confusions (83 →47), alongside smaller reductions in Neutral →Positive (45 →36) and Neutral →Negative (87 →80). In contrast, adjacent errors remained the dominant failure mode in both conditions, reflecting intrinsic boundary ambiguity between Negative and Neutral discourse in cancer support narratives.

To estimate irreducible difficulty, we identified test instances misclassified by *all* evaluated architectures (Logistic Regression, Random Forest, LightGBM, GRU, and ALBERT). We observed 127 such cases (6.1%) in the regular condition and 142 (6.8%) in the augmented condition, with 103 instances failing universally in both. These global-failure cases indicate a non-trivial subset of posts whose sentiment remains intrinsically ambiguous or context-dependent even after contextual modeling and token-based augmentation, reinforcing that deployment should be framed as *decision support* with human review—particularly for low-confidence or high-impact predictions.

Overall, augmentation primarily improved performance by (i) reducing the most consequential polarity reversals for ALBERT and (ii) correcting a larger number of sequential-model errors for GRU, while leaving adjacent boundary ambiguity as the dominant residual challenge. This pattern suggests that future improvements should focus less on longer-context capacity and more on boundary-specific strategies (e.g., calibration, class-aware thresholds, and targeted labeling of borderline Negative/Neutral instances) to reduce systematic boundary drift without sacrificing sensitivity to high-need content.

## 4 Discussion

This study evaluated emotional tone classification for posts in online cancer peer-support communities under two substantive methodological extensions: (i) supervision derived from an LLM-based annotation protocol (AI labels) and (ii) token-based incorporation of LLM-extracted structured context (reporter role and cancer type) as additional model input. Across a common experimental pipeline and a held-out test set, we observed two overarching findings. First, the LLM annotator induced a marked distributional shift relative to the dataset’s original labels, particularly by reallocating a large share of posts toward the *very negative* category, and this shift remained (though attenuated) after collapsing to a three-class modeling target. Second, despite this labeling shift, token-based augmentation provided consistent improvements in discriminative performance across diverse model families, with the largest gains for lower-capacity sequential modeling (GRU) and measurable improvements even for a fine-tuned transformer (ALBERT).

### LLM labeling shift: interpretability and implications

The most salient descriptive result is the divergence between human labels and LLM-derived labels in the four-class scheme. The LLM assigned nearly half of all posts to *very negative*, substantially increasing the prevalence of severe distress language relative to the original labels. This pattern is consistent with a systematic difference in intensity thresholds: the LLM appears to treat a wider set of worry, uncertainty, symptom narratives, and treatment-related difficulties as “very negative” rather than reserving that label for extreme affect. Prior work has emphasized that LLMs can behave as confident but systematically shifted raters depending on rubric framing and latent norms learned during pretraining [11, 12]. From a measurement standpoint, this shift implies that AI labels should not be interpreted as a drop-in replacement for human labels without explicit calibration or auditing, particularly when analyses depend on absolute prevalence of distress categories.

Importantly, the distributional divergence was reduced under the three-class collapse used for modeling (Negative/Neutral/Positive), suggesting that much of the disagreement arises within the negative subspace. This supports the practical motivation for collapsing adjacent negative categories when the downstream goal is screening rather than fine-grained affect intensity estimation. However, even under the collapsed target, the LLM assigned substantially more posts to Negative and fewer to Neutral/Positive, which may bias downstream summaries of community emotional tone toward higher distress. In applied settings, this could have both benefits and risks: a conservative rater may improve sensitivity to potentially high-need content, but may also increase false alarms and dilute prioritization capacity if operational triage thresholds are not adjusted.

These findings reinforce broader concerns about label noise, disagreement, and learnability in subjective text annotation [8, 9]. When supervision is produced by an LLM, label shift becomes a first-order methodological artifact. A practical implication is that evaluations should (i) report label distribution comparisons, (ii) quantify uncertainty and robustness to labeling choice, and (iii) frame model outputs as *decision support* rather than ground truth, especially for high-impact use cases in mental health and supportive oncology [5, 7].

### Token-based augmentation as lightweight context integration

Despite labeling shift, augmentation with LLM-extracted role and cancer-type tokens improved performance consistently across all model families. The uniform direction of gains across TF–IDF baselines, gradient boosting, GRU, and ALBERT suggests that these coarse context features provide complementary signal beyond lexical content alone. Conceptually, reporter role can change the semantics of negative language (e.g., caregiving burden vs. patient symptom distress), and cancer type may correlate with treatment trajectories and symptom narratives that shape tone expression. Notably, these structured variables were extracted from the same input text, so they do not add external information; rather, they function as an upstream information-extraction layer that makes salient context explicit to the classifier.

The largest improvements occurred for the GRU model, which also had the lowest baseline performance. This is consistent with prefix tokens acting as high-salience anchors for a lower-capacity sequential encoder, helping it resolve ambiguity early in the sequence.

For ALBERT, gains were smaller in magnitude but still consistent, indicating that even transformer-based encoders can benefit from explicit context markers that reduce the burden of implicitly inferring role/type from noisy and variable text. In practice, token augmentation is attractive because it is model-agnostic, requires minimal architectural change, and can be integrated into existing text pipelines with little engineering overhead.

Class-wise results for ALBERT further suggest that augmentation altered decision boundaries in operationally meaningful ways. Improvements in Negative performance indicate reduced confusion for clearly distressed narratives, while changes for Neutral and Positive reveal a nuanced trade-off: augmentation increased Neutral recall (recovering more neutral posts) while slightly reducing precision, and improved Positive precision at some cost to recall. This pattern is compatible with augmentation helping the model become more selective about minority-class assignments, particularly for Positive where false positives can be common under imbalance. From a screening perspective, improved Positive precision may better preserve the interpretability of “positive coping” detection, while Neutral recall gains may reduce over-triage of informational posts as negative.

### Error structure and operational consequences

The ordinal-aware error analysis provides an additional perspective beyond aggregate metrics. Adjacent-category confusions remained the dominant error mode, reflecting inherent boundary ambiguity between Negative and Neutral discourse in cancer support narratives. However, augmentation reduced the most consequential polarity-reversing mistakes for ALBERT (Negative ↔ Positive), indicating that explicit context tokens can help avoid extreme misinterpretations that would be problematic in real-world triage or monitoring. The persistence of adjacent confusions suggests that future improvements may be better targeted toward boundary-sensitive strategies—for example, calibration and selective abstention, class-aware thresholds, or additional rubric-guided labeling for borderline Negative/Neutral instances.

The subset of instances misclassified by all model families highlights another practical consideration: a non-trivial fraction of posts may be intrinsically ambiguous or require context not present in the text (e.g., prior thread history, temporal trajectory, or user baseline). This observation aligns with broader findings in mental-health NLP that model performance is often limited by construct ambiguity and annotation variability rather than solely by model capacity [5, 6]. For deployment, this supports a workflow where low-confidence cases are routed to human review rather than forcing a single-label decision.

### Model comparisons and the role of modern encoders

Across both regular and augmented conditions, ALBERT achieved the strongest overall discrimination, particularly in macro OvR AUC, consistent with established advantages of pretrained transformers for sentence-level classification [14, 15]. Nevertheless, classical TF–IDF baselines performed competitively in weighted F1, with overlapping confidence intervals among top models. This has practical implications: in settings where computational resources, latency, or interpretability constraints favor simpler models, TF–IDF baselines may provide strong performance, and augmentation can further narrow gaps. Conversely, when the goal is robust discrimination across classes (as reflected by macro AUC), transformers may provide additional benefit.

### Limitations

This work has several limitations. First, the LLM annotator was applied in a single configuration, and labeling behavior may vary with prompt design, temperature, model version, or rubric specificity. Systematic prompt ablations and multi-annotator consensus protocols could help disentangle whether the observed severity shift is intrinsic to the model or a consequence of instruction framing. Second, auxiliary variables were restricted to two coarse attributes; additional structured factors (e.g., treatment stage, symptom mentions, prognosis uncertainty) could further improve classification but may be harder to extract reliably and may raise additional ethical concerns. Third, while we used a held-out test set with bootstrapped confidence intervals and paired statistical tests, the results are specific to this dataset and to the three-class collapse; external validation on other cancer community corpora would strengthen generalizability [13]. Finally, posts are de-identified, but sentiment modeling in health-related communities raises ethical risks, including misclassification harms, privacy concerns, and downstream misuse; such models should be deployed only with clear governance, transparency, and human oversight [20, 21].

### Future directions

Several directions follow from these findings. First, future work should evaluate *calibration* and uncertainty-aware decision support, including selective abstention on borderline posts and evaluation of probability quality (e.g., with proper scoring rules) to better support triage. Second, rubric-guided or multi-pass LLM annotation could be used to reduce intensity threshold drift and to quantify annotator uncertainty, potentially improving label consistency and downstream learnability. Third, beyond token augmentation, more structured integration approaches (e.g., multi-task learning that jointly predicts tone and extracted attributes) may improve boundary resolution, though at greater implementation complexity. Finally, external validation across platforms and communities is essential to assess robustness to domain shift and to mitigate biases stemming from platform-specific language norms.

## Conclusion

In summary, LLM-based annotation substantially reshaped label distributions for emotional tone in cancer peer-support posts, emphasizing the need to treat AI labels as a distinct measurement instrument rather than a neutral substitute for human judgment. At the same time, LLM-extracted structured context, incorporated via simple token-based augmentation, improved predictive performance consistently across model families and reduced the most severe ordinal errors for the best-performing transformer. These results suggest a pragmatic hybrid approach: use LLMs not only as annotators but also as lightweight context extractors, while explicitly auditing label shift and maintaining human oversight for ambiguous or high-impact cases.

## Data Availability

The original dataset is publicly available on Kaggle: https://www.kaggle.com/datasets/irinhoque/mental-health-insights-vulnerable-cancer-patients Annotated dataset will be provided upon request during review

